# Prevalence and Clinical Characteristics of Patients with Ischemic Stroke with *JAK2* V617F Mutation and Normal Blood Counts

**DOI:** 10.64898/2026.05.01.26352265

**Authors:** Toshiyuki Hayashi, Takashi Shimoyama, Yasuhiro Nishiyama, Hiroki Yamaguchi, Takehiro Katano, Yuki Sakamoto, Satoshi Suda, Kazumi Kimura

## Abstract

**Objective:** The *JAK2* V617F mutation increases the risk of thrombosis in patients with myeloproliferative neoplasms (MPNs). However, it remains unclear whether individuals who carry the *JAK2* V617F mutation without MPN also have an increased risk of stroke.

**Methods:** We prospectively tested for the *JAK2* 617F mutation in consecutive patients with acute ischemic stroke or transient ischemic attack (TIA) admitted between January 2020 and September 2024. Patients with overt MPN or abnormal blood counts were excluded. We used allele-specific PCR to detect the mutations.

**Results:** In total, 921 patients (median age, 77 years; 557 men (62%); TIA, 32 patients) were enrolled in this study. Among them, 11 patients (1.2%; median age, 72 years; 8 male) tested positive for the *JAK2* V617F mutation. There were no significant differences in clinical background, including age, sex, BMI, comorbidities, or history of thrombosis, between the positive and negative groups. The blood count and coagulation parameters did not differ significantly between the two groups. Among the 11 patients in the positive group, 9 had embolic stroke and 2 had thrombotic stroke. Embolic stroke of undetermined source (ESUS) was more frequently observed in the positive group than in the negative group (45 vs. 13%; p=0.002). Stroke severity and outcomes did not differ between the two groups.

**Discussion:** Approximately 1% of patients with acute ischemic stroke or TIA carry the *JAK2* V617F mutation despite normal blood counts. Of the 11 mutation-positive patients, nine (82%) exhibited embolic imaging features and five (45%) met the ESUS criteria, whereas other clinical characteristics did not differ significantly from the mutation-negative group.

## Introduction

Polycythemia vera (PV) and essential thrombocythemia (ET) are subtypes of myeloproliferative neoplasms (MPN) characterized by chronic clonal proliferation of myeloid cells due to hematopoietic stem cell abnormalities. The Janus Kinase 2 (JAK2) valine 617 phenylalanine (V617F) mutation is detected in the majority of patients with MPN, in 95% of patients with PV, and in half or more of those with ET.^1^ Approximately 30% of patients with MPN develop thrombosis,^2, 3^ and a positive *JAK2* V617F mutation is thought to be a risk factor for thrombotic events.^4-6^

Ischemic stroke in patients with MPN presents with a wide spectrum of clinical manifestations. Previous studies have reported frequent large vessel stenosis or occlusion,^7^ whereas others have reported multiple cerebral infarcts^8^ or lacunar lesions^9^. Several reports have indicated that most MPN-related ischemic strokes have favorable outcomes; however, similar to conventional stroke, recurrence has also been reported.

Recently, clonal hematopoiesis has been reported in individuals with normal blood counts.^10, 11^ Individuals with clonal hematopoiesis are known to be at an increased risk of developing hematologic malignancies.^10, 11^ Some studies have reported that patients with ischemic stroke are *JAK2* V617F-positive despite having normal blood counts, and a proportion of those individuals were subsequently diagnosed with MPN.^12-14^ *JAK2* V617F positivity itself may increase the risk of stroke, particularly in cases of embolic stroke of undetermined source (ESUS). However, in patients with normal blood counts, the prevalence and clinical characteristics of ischemic stroke associated with *JAK2* V617F mutation remain unclear.

This study aimed to investigate the prevalence and clinical characteristics of patients with ischemic stroke who were *JAK2* V617F-positive despite normal blood counts and without MPN.

## Methods

### Study design and participants

We conducted a prospective study from January 2020 to September 2024 at Nippon Medical School Hospital. The trial is registered in the UMIN Clinical Trials Registry (registration number: UMIN000047136). The inclusion criteria for this study were as follows: (1) patients with ischemic stroke or transient ischemic attack (TIA); (2) patients admitted within 7 days of stroke onset; (3) patients without elevated blood cell counts suggestive of MPN; (4) patients, family, or other representatives consented to participate in the study; and (5) patients aged 18 years or older. The exclusion criteria were as follows: (1) patients who did not consent to participate in this study, (2) patients for whom serum to measure *JAK2* V617F mutation was not available, and (3) patients with known MPN and/or the presence of *JAK2* V617F mutation.

### Clinical examination

Clinical characteristics included sex, age, premorbid modified Rankin scale (mRS) score, and cardiovascular risk factors, such as hypertension (HT), dyslipidemia (DL), diabetes mellitus, chronic kidney disease, congestive heart failure, smoking, and alcohol consumption. Information on the direct oral anticoagulant dose, history of stroke, and history of vascular disease, including peripheral arterial disease, aortic plaque, and acute cardiac syndrome, was also obtained. Stroke severity on admission and discharge was assessed using the National Institutes of Health Stroke Scale (NIHSS) and mRS scores, respectively. Routine blood biochemical analyses were performed on admission. A hypercytosis in blood cells was determined using the international consensus classification of PV and ET^15^ as follows: 1) hemoglobin/hematocrit (Hb/Hct) concentrations above 16.5 g/dL/49% in men and 16 g/dL/48% in women, or 2) platelet count ≥450×10^9^/L. The patient underwent magnetic resonance imaging (MRI) on admission to diagnose acute ischemic stroke. The site of arterial occlusion was determined using magnetic resonance angiography. Gradient-recalled echo T2*-weighted images were assessed for evidence of cerebral microbleeds (defined as parenchymal hemorrhages ≤10 mm in diameter).^16^ Patients with a contraindication to MRI were evaluated on computed tomography angiography or conventional angiography. Stroke etiology, including small vessel occlusion, large artery atherosclerosis, and cardioembolism, were defined using the Stop Stroke Study Trial of Org 10720 in Acute Stroke Treatment classification.^17^ Stroke mechanisms were classified as thrombotic, embolic and hemodynamic based on the Classification of Cerebrovascular Diseases III published by National Institute of Neurological Disorders and Stroke.^18^ The criteria for TIA and ESUS were based on criteria proposed by the Cryptogenic Stroke/ESUS International Working Group^19^ and American Heart Association/American Stroke Association Stroke Council^20^, respectively.

### Measurement of *JAK2* V617F mutation

Consecutive patients admitted during the study period were tested for *JAK2* V617F mutation. Mononuclear cells from the peripheral blood were isolated by density gradient centrifugation using lymphocyte separation medium (Organon, Durham, NC, USA). Genomic DNA was extracted using a QIAamp DNA Mini Kit (Qiagen, Hilden, Germany) according to the manufacturer’s protocol. The DNA concentration was determined by Nano Drop^®^ at a wavelength of 260, and the absorption ratio of a pure sample DNA at a wavelength of 260 to 280 nm was calculated (260/280 ratio). Allele-specific polymerase chain reaction (PCR) for *JAK2* genotyping was performed using 20 ng genomic DNA, 45 nM forward primer *JAK2*-F, and 22.5 nM each of the allele-specific reverse primers *JAK2*-R-T and *JAK2*-R-G. The following primers were used: 5′-CTGAATAGTCCTACAGTGTTTTCAGTTTCA-3′ (forward primer), 5′-AGCATTTGGTTTTAAATTATGGAGTATATT-3′ (reverse primer), and 5′-ATCTATAGTCATGCTGAAAGTAGGAGAAAG-3′ (mutant primer). Thirty PCR cycles with denaturing at 94°C for 30 seconds, annealing at 61°C for 30 seconds, and extension at 72°C for 30 seconds were applied. PCR products were analyzed by agarose gel electrophoresis and ethidium bromide staining. The forward and mutant primers amplify a 364 bp product (both mutant and wild-type alleles), whereas the forward and reverse primers amplify a 203 bp product (when the patient carried the *JAK2* V617F mutation).

### Statistical analysis

First, we investigated the proportion of patients who tested positive for *JAK2* V617F mutation. Next, patients were divided into two groups: those who tested positive for the *JAK2* V617F mutation (positive group) and those who tested negative (negative group). Clinical, laboratory, and radiographic characteristics were compared between the two groups. Data are presented as median (interquartile range [IQR]) or number (%). Intergroup differences were assessed using the chi-square test or Wilcoxon rank-sum test, as appropriate. All analyses were performed using JMP 13 statistical software (SAS Institute Inc., Cary, NC, USA). The level of statistical significance was set at p < 0.05.

### Ethical considerations

The Nippon Medical School Ethics Review Committee approved the study protocol (approval number: B-2020–150). Written informed consent was obtained from all the patients whose data were included in the study.

## Results

Between January 2020 and September 2024, 1,009 patients with cerebral infarction or TIA were hospitalized (Figure 1). After excluding patients whose blood samples could not be tested due to COVID-19 co-infection (n=8), those who did not consent to study participation (n=76), and those already known to be *JAK2* V617F mutation-positive (n=4), 921 patients underwent testing for the *JAK2* V617F mutation. Furthermore, as mononuclear cell isolation failed in 5 patients, 916 patients were included in the final analysis. Among these, 11 patients were confirmed to be positive for the *JAK2* V617F mutation. Therefore, the proportion of patients who tested positive for the *JAK2* V617F mutation was 1.2%.

**Figure 1.**
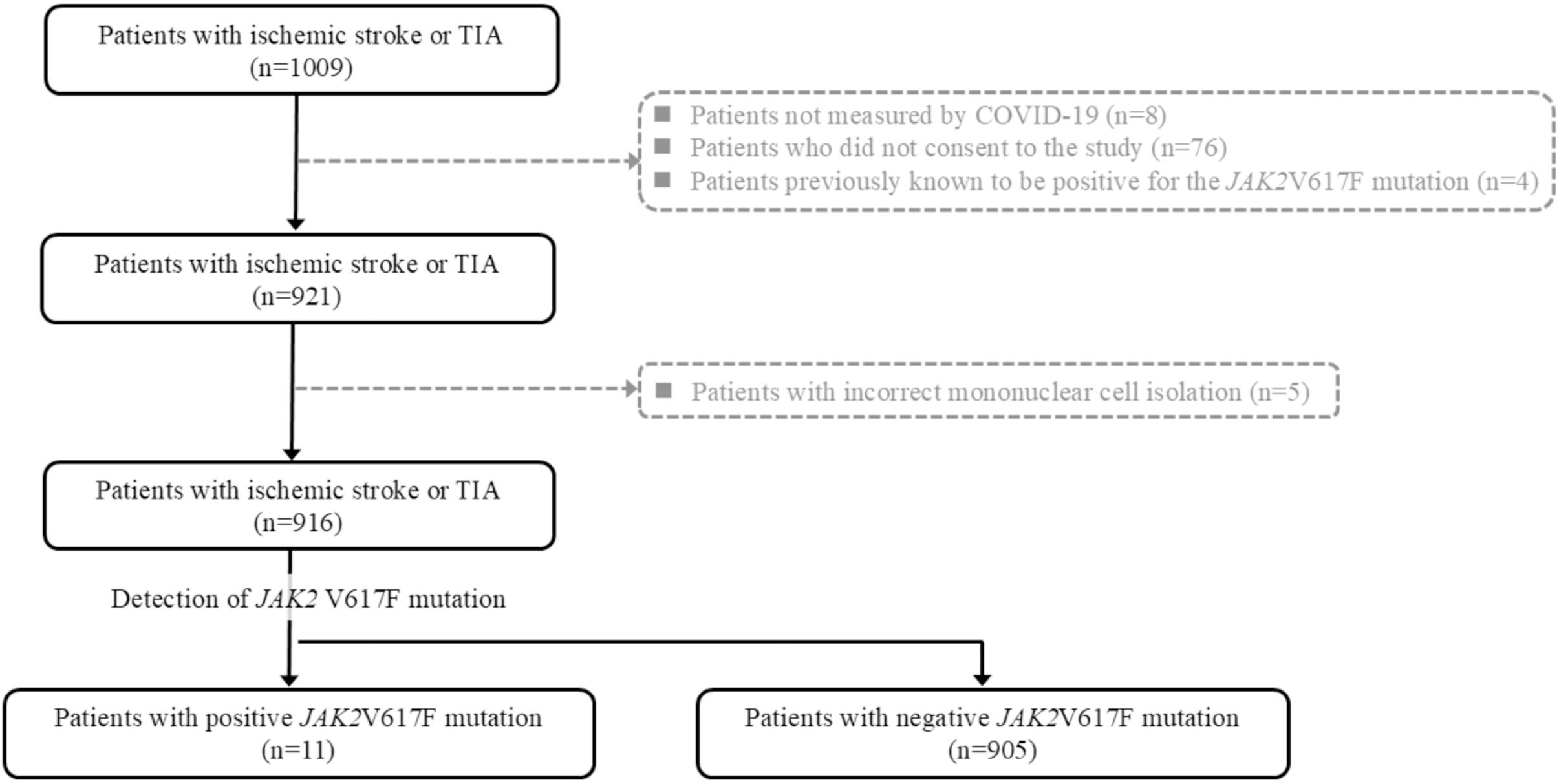
Study Flow Chart During the study period, 1,009 patients were hospitalized for cerebral infarction or transient ischemic attack (TIA). Blood samples from 921 patients were tested for the *JAK2* V617F mutation and 11 were found to be positive.

**Figure 2.**
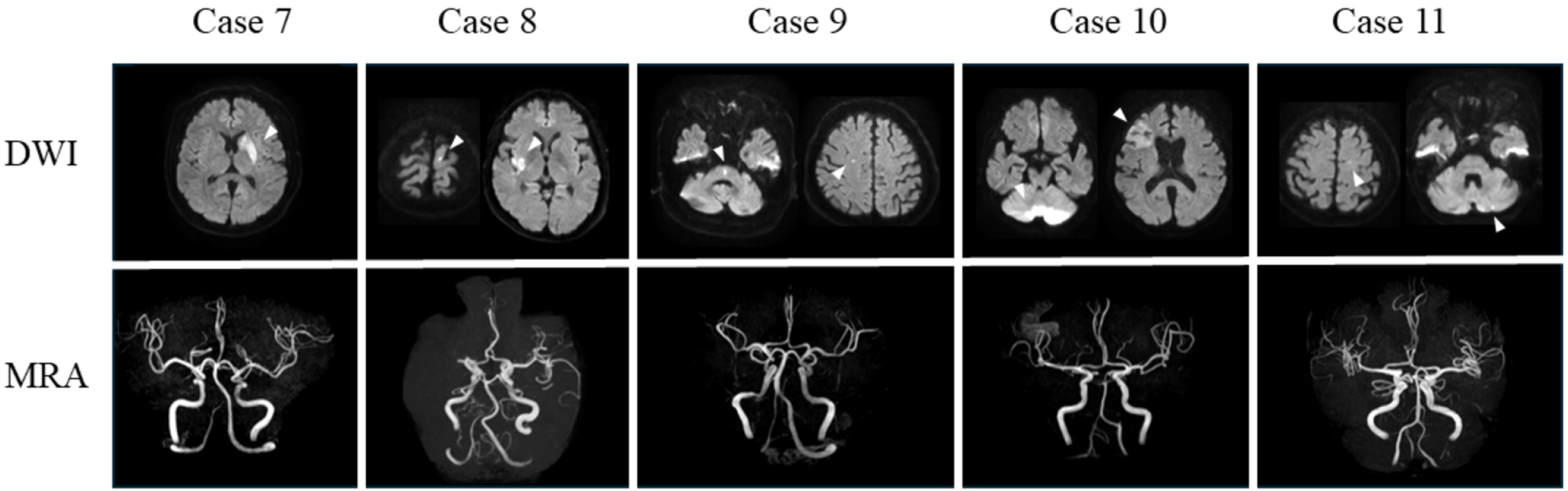
Cases with positive *JAK2* V617F mutation fulfilling ESUS diagnostic criteria Among *JAK2* V617F mutation-positive cerebral infarctions with normal blood counts, five met the diagnostic criteria for embolic stroke of undetermined source (ESUS). The arrowhead indicates the cerebral infarction location.

Age, sex, BMI, comorbidities, and history of thrombosis did not differ significantly between the two groups (Table 1). Stroke severity, antithrombotic therapy, and clinical outcomes were comparable, with no significant intergroup differences. Blood counts, coagulation profiles, and BNP levels did not differ between the two groups (Table 2). There was also no difference between the two groups in the presence of multiple infarct locations or arterial occlusions. Patients classified as having TIA, small vessel occlusion, large artery atherosclerosis, cardioembolism, or other etiologies did not differ significantly between the two groups. In contrast, the number of patients who met the ESUS criteria was significantly higher in the positive group than in the negative group (45 vs. 13%; p=0.002). Of the 122 patients classified as having ESUS, 5 (4%) tested positive for the *JAK2* V617F mutation.

**Table 1.** Clinical characteristics.

| Variable | *JAK2* V617F mutation | *JAK2* V617F mutation | p |
| --- | --- | --- | --- |
|  | positive group | negative group |  |
|  | n = 11 | n = 905 |  |
| Age, years, median (IQR) | 72 (63-78) | 77 (67-84) | 0.55 |
| Male, n (%) | 8 (73) | 561 (62) | 0.47 |
| BMI, median (IQR) | 21 (19-27) | 23 (20-25) | 0.59 |
| Comorbidity |  |  |  |
| HT, n (%) | 7 (64) | 425 (48) | 0.58 |
| DL, n (%) | 7 (64) | 407 (45) | 0.22 |
| DM, n (%) | 3 (27) | 226 (25) | 0.87 |
| CKD, n (%) | 0 (0) | 81 (1) | 0.29 |
| AF, n (%) | 2 (18) | 227 (25) | 0.94 |
| Congestive heart failure, n (%) | 0 (0) | 81 (1) | 0.29 |
| Tumors, n (%) | 2 (18) | 109 (12) | 0.74 |
| Smoking, n (%) | 6 (55) | 489 (54) | 0.93 |
| Alcohol, n (%) | 4 (36) | 407 (45) | 0.56 |
| History of stroke, n (%) | 2 (18) | 163 (18) | 0.99 |
| History of ischemic heart disease, n (%) | 0 (0) | 63 (7) | 0.38 |
| Pre-admission antiplatelet use, n (%) | 2 (18) | 172 (20) | 0.39 |
| Pre-admission anticoagulant use, n (%) | 1 (9) | 109 (12) | 0.42 |
| Preadmission mRS, median (IQR) | 0 (0-2) | 0 (0-1) | 0.90 |
| NIHSS at admission, median (IQR) | 3 (2-11) | 3 (1-9) | 0.44 |
| Treatment |  |  |  |
| rt-PA, n (%) | 3 (27) | 118 (12) | 0.14 |
| Endovascular therapy, n (%) | 2 (18) | 154 (17) | 0.96 |
| Antiplatelet therapy, n (%) | 8 (73) | 543 (60) | 0.82 |
| Anticoagulation therapy, n (%) | 3 (27) | 281 (32) | 0.91 |
| NIHSS at discharge, median (IQR) | 0 (0-2) | 0 (0-1) | 0.55 |
| mRS at discharge, median (IQR) | 2 (1-4) | 2 (0-4) | 0.33 |
| Death, n (%) | 0 (0) | 25 (3) | 0.57 |

AF, atrial fibrillation; BMI, body mass index; CKD, chronic kidney disease; DL, dyslipidemia; DM, diabetes mellites; IQR, interquartile range; HT, hypertension; mRS, modified Rankin Scale; NIHSS, National Institutes of Health stroke scale; rt-PA, recombinant tissue-type plasminogen activator.

**Table 2.** Laboratory findings in *JAK2* V617F mutation positive and negative group.

| Variable | *JAK2* V617F mutation<br>positive group | *JAK2* V617F mutation<br>negative group | p |
| --- | --- | --- | --- |
|  | n = 11 | n = 905 |  |
| WBC, $\times 10^3/\mu\text{L}$, median (IQR) | 6.2 (5.3-7.6) | 6.8 (5.5-8.6) | 0.25 |
| Hemoglobin, g/dl, median (IQR) | 12.8 (10.6-14.7) | 13.1 (11.8-14.4) | 0.72 |
| Platelet count, $\times 10^3/\mu\text{L}$, median (IQR) | 231 (182-285) | 210 (171-254) | 0.23 |
| D-dimer, $\mu\text{g/ml}$, median (IQR) | 1.2 (0.6-5.3) | 1.1 (0.7-2.1) | 0.71 |
| BNP, pg/ml, median (IQR) | 56.2 (10.6-122.7) | 46.0 (16.4-140.9) | 0.53 |

BNP, brain natriuretic peptide; IQR, interquartile range; WBC, white blood cell

**Table 3.** Radiographic findings in *JAK2* V617F mutation positive and negative group.

|  | *JAK2* V617F mutation | *JAK2* V617F mutation |  |
| --- | --- | --- | --- |
| Variable | positive group | negative group | p |
|  | n = 11 | n = 905 |  |
| Stroke etiology |  |  |  |
| TIA, n (%) | 0 (0) | 35 (4) | 0.84 |
| Small vessel occlusion, n (%) | 2 (18) | 141 (15) | 0.78 |
| Large artery atherosclerosis, n (%) | 2 (18) | 148 (16) | 0.88 |
| Cardioembolism, n (%) | 2 (18) | 245 (27) | 0.53 |
| Others or undetermined, n (%) | 5 (45) | 348 (38) | 0.60 |
| ESUS, n (%) | 5 (45) | 117 (13) | 0.002 |
| Stroke mechanism |  |  |  |
| Thrombotic, n (%) | 2 (18) | 309 (34) | 0.27 |
| Embolic, n (%) | 9 (82) | 573 (63) | 0.19 |
| Hemodynamic, n (%) | 0 (0) | 30 (3) | 0.54 |
| Multiple ischemic lesion | 6 (55) | 285 (31) | 0.10 |
| Arterial occlusion at admission, n (%) | 4 (36) | 337 (37) | 0.97 |
| Arterial stenosis at admission, n (%) | 3 (27) | 291 (32) | 0.63 |

ESUS, embolic stroke of undetermined source; TIA, transient ischemic attack

**Table 4-1.** Clinical characteristics of patients with positive *JAK2* V617F mutation.

| Case | Age/Sex | Vascular risk factors | History or comorbidity of vascular event | Comorbidity of Cancer | Stroke etiology | Stroke mechanism | Single/multiple lesion | Arterial stenosis | Arterial occlusion |
| --- | --- | --- | --- | --- | --- | --- | --- | --- | --- |
| 1 | 70s/F | HT | Deep vein thrombosis | - | Large artery atherosclerosis | Embolic | Single | M1 | M1 |
| 2 | 90s/M | HT | - | - | Large artery atherosclerosis | Embolic | Multiple | BA | - |
| 3 | 60s/M | HT, DL, DM, Smoking | - | - | Small vessel occlusion | Thrombotic | Single | - | - |
| 4 | 70s/M | HT, DL, Smoking | - | - | Small vessel occlusion | Thrombotic | Single | - | - |
| 5 | 70s/M | AF | - | - | Cardioembolism | Embolic | Multiple | - | M2 |
| 6 | 70s/M | HT, DL, AF, Smoking | - | + | Cardioembolism | Embolic | Multiple | ICA | M2 |
| 7 | 40s/M | HT, DL, DM | History of TIA | - | Others (ESUS) | Embolic | Single | - | - |
| 8 | 50s/M | HT, Smoking | - | - | Others (ESUS) | Embolic | Single | - | M1 |
| 9 | 70s/M | DL, DM, Smoking | History of ischemic stroke | - | Others (ESUS) | Embolic | Multiple | - | - |
| 10 | 70s/F | Smoking | Deep vein thrombosis | + | Others (ESUS) | Embolic | Multiple | - | - |
| 11 | 90s/F | DL | - | - | Others (ESUS) | Embolic | Multiple | - | - |

**Table 4-2.**
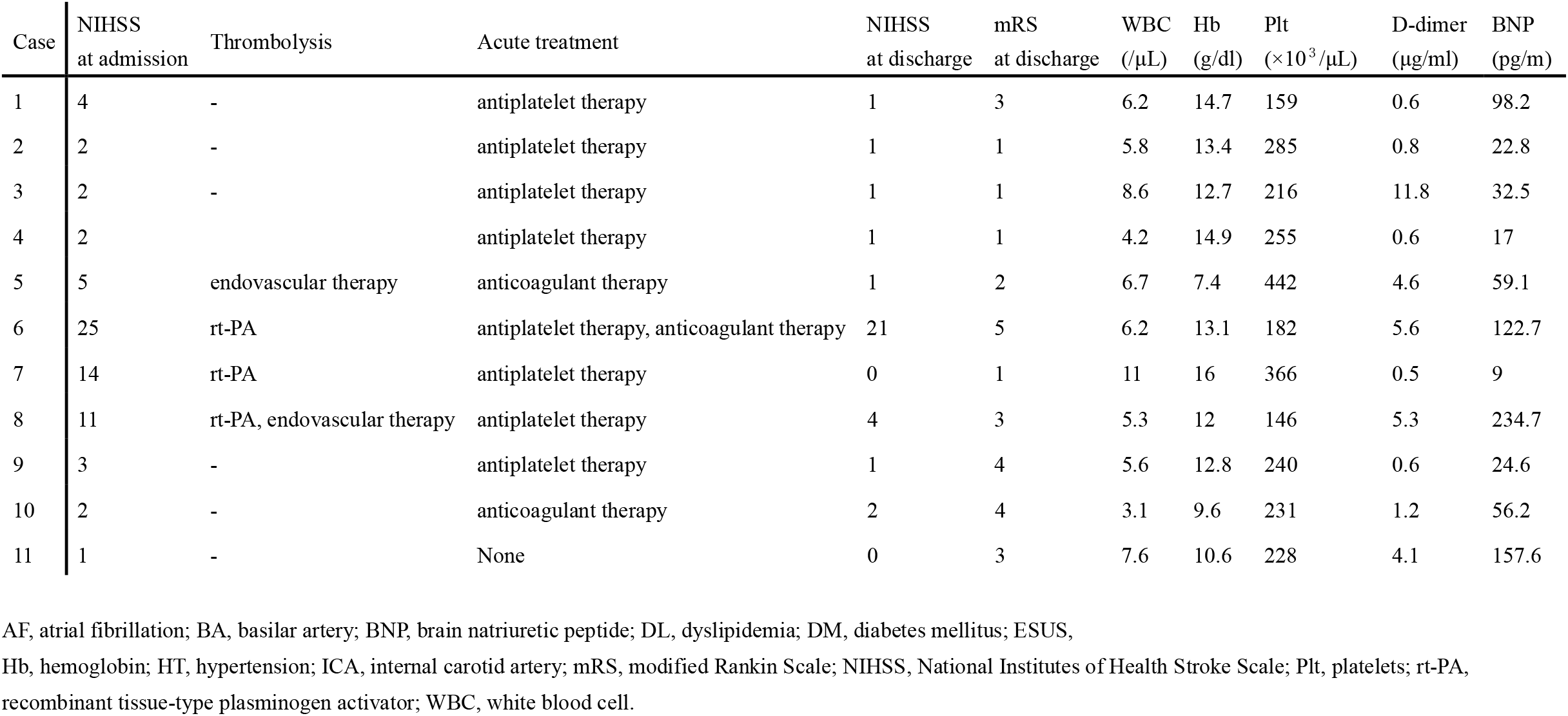
Clinical characteristics of patients with positive *JAK2 V617F* mutation.

Table 4 summarizes 11 patients (median age, 72 years; 8 males) hospitalized for ischemic stroke who were positive for the *JAK2* V617F mutation with normal blood counts. Vascular risk factors included HT in seven cases (64%), DL in six cases (55%), and smoking in 6 (55%). Two (18 %) patients had a history of ischemic stroke or TIA. Comorbid cancer was present in two patients (18%). Regarding the stroke mechanism, embolic stroke was most frequently observed, occurring in 9 patients (82%), whereas thrombotic stroke was identified in 2 patients (18%). Regarding stroke subtype, others or undetermined cases were most frequently observed in 5 cases (45%), all of whom met the criteria for ESUS. Arterial stenosis and occlusion were observed in 3 (27%) and 4 (36%) patients, respectively. Of these, 2 patients (18%) underwent mechanical thrombectomy. Eight (73%) patients received antiplatelet therapy. Ten patients (91%) were discharged with an NIHSS score <5, but only 5 patients (45%) achieved a favorable outcome of mRS 0-2 at discharge due to advanced age and low pre-admission ADL levels.

## Discussion

This study examined the prevalence and clinical characteristics of ischemic stroke patients with *JAK2* V617F mutation and normal blood counts. Our results showed that approximately 1% of patients were positive for the *JAK2* V617F mutation. Among the 11 positive patients, nine (82%) demonstrated radiographic features of embolism, and five (45%) met the diagnostic criteria for ESUS.

The prevalence of the *JAK2* V617F mutation in patients with ischemic stroke has been reported to be 0.7–11.3%.^13, 21-24^ One reason for the variation in positive rates is thought to be the difference in the detection sensitivity of *JAK2* V617F mutation. In the present study, allele-specific PCR was used to detect the mutations. Previous studies^13, 23^ using the same method reported positivity rates of 0.7–1.1%, which is consistent with the results of this study. However, studies using methods that can detect mutations at an allele burden of 0.1% or lower showed positive rates as high as 2.5– 11.3%.^21, 22, 24^ In addition, the inclusion of patients with MPN in these studies^21, 22, 24^ may also contribute to this high detection rate. Our study focused on patients with ischemic stroke, excluding those with overt MPN or elevated blood counts, which is considered a unique feature of this study. Furthermore, the *JAK2* V617F mutation positivity rate may vary among population groups. Kristiansen et al.^22^ reported the highest frequency of *JAK2* V617F mutation positivity in 11.3% of patients with ischemic stroke and also found positivity in 3.8% of the general population in Denmark. Further studies comparing the prevalence of *JAK2* V617F mutation across racial groups are required.

In this study, there were no significant differences in the clinical backgrounds of the patients with and without *JAK2* V617F mutations. This was also consistent with previous reports of mutation-positive patients with cerebral venous thrombosis (CVT).^25^ Studies investigating the *JAK2* V617F mutation in patients with CVT presenting with normal blood counts have reported that patients with positive mutations tend to be older than patients with negative mutation.^26^ In this study, 8 out of 11 mutation-positive cases (73%) were aged 70 years or older. However, in the present study, there was no difference in age between the two groups, as most patients were elderly at onset.

Of the 11 patients with a positive *JAK2* V617F mutation, 5 (45%) met the diagnosis of ESUS. To the best of our knowledge, no studies have investigated the association between ESUS and *JAK2* V617F mutation. Among patients with a history of MPN who developed ischemic stroke, 13–18% experienced major artery occlusion.^8, 12^ Furthermore, another study reported patients with ischemic stroke in the watershed area without artery stenosis.^27^ Case 8 in this study presented M1 occlusion; however, routine investigations could not identify the source of the embolism. In this case, positive *JAK2* V617F mutation might be considered to be associated with the occurrence of embolic stroke.

As a mechanism for arterial occlusion, it is hypothesized that patients with MPN form mural thrombi, potentially acting as an embolus.^28-30^ The present study found that 3 out of 11 (27%) patients in the *JAK2* V617F mutation-positive group had major artery stenosis. Cases 1 and 2 in this study were both elderly and had HT but no other risk factors for atherosclerosis despite presenting with severe stenosis of the major artery. The *JAK2* V617F mutation may be associated with this finding. Another study reported that ATBI was the most common subtype among patients positive for *JAK2* F617F mutation.^24^

Among our patients with the *JAK2* V617F mutation, future monitoring of blood tests may reveal rising blood cell counts, leading to a diagnosis of MPN. However, some patients did not develop elevated cell counts. Further research is required in this area.

This study had some limitations. First, there is a potential for selection bias in single-center studies. This study focused on Japanese participants and did not examine the differences across racial groups. Second, a relatively high proportion of patients with severe cerebral infarction or those without family members were unable to provide consent, suggesting the possibility of selection bias. Third, the low prevalence of the JAK2V617F mutation made it difficult to perform a statistical analysis of its characteristics. Furthermore, we were unable to measure the allele burden of the *JAK2* V617F mutation. However, the relationship between allele burden and clinical characteristics has not yet been examined.

In conclusion, *JAK2* V617F mutation was detected in approximately 1% of patients with acute ischemic stroke/TIA without overt MPN and with normal blood counts; mutation-positive patients fulfilled the ESUS criteria more frequently.

## Data Availability

Data are available from the corresponding author upon reasonable request.

## Non-standard Abbreviations and Acronyms

CVT: cerebral venous thrombosis
DL: dyslipidemia
ESUS: embolic stroke of undetermined source
ET: essential thrombocythemia
HT: hypertension
*JAK2* V617F: Janus Kinase 2 valine 617 phenylalanine
MPN: myeloproliferative neoplasms
mRS: modified Rankin scale
NIHSS: National Institutes of Health Stroke Scale
PCR: polymerase chain reaction
PV: polycythemia vera
TIA: transient ischemic attack.

## Acknowledgments

We are grateful to Miho Miyata, Kunihito Arai, and Akiko Morijiri for their significant contributions in detecting the *JAK2* V617F mutation.

